# Biological, biomechanical, and pain sensitivity effects of walk-run in people with self-reported knee osteoarthritis

**DOI:** 10.1101/2025.10.16.25338143

**Authors:** Tsun (Tim) Chow, Nasir Uddin, Christopher McManus, Zainab Altai, Qichang Mei, David W. Evans, Sally Waterworth, Bernard X.W. Liew

## Abstract

**Objectives:** Participating in small volumes of high-impact exercises in older adults, like running, is beneficial to mitigate age-related musculoskeletal deterioration. The biological and pain effects of small volumes of running integrated within a walking program in people with knee osteoarthritis (OA) are unclear. This study aims to investigate the biological and pain responses to a walk-run program in people with self-reported knee OA.

**Methods:** Eight participants with self-reported knee OA completed a 25-minute walk-run program on a force-plate instrumented treadmill with optical motion capture. Peak knee moment and cumulative knee moment impulse were quantified during this exercise task. Pressure pain threshold (PPT) was measured at the knee and elbow before and immediately after the task. A cartilage stress marker, cartilage oligomeric matrix protein (COMP), was measured at baseline, immediately after exercise (Post) and 30 minutes after exercise (Post30).

**Results:** PPT did not significantly change at the knee (Δ7.28%, *P* = 0.700) or elbow (Δ-9.15%, *P* = 0.496). Percentage COMP significantly increased from baseline immediately post-exercise (Hedges’ *g* = 1.43), then returned to baseline concentration at Post30 (Hedges’ *g* = 0.07). No significant correlations were observed between COMP and knee joint moment indices.

**Conclusions:** Including short periods of running within a primarily walking-based session does not lead to prolonged elevations in cartilage stress markers or altered pain sensitivity. Walk-run may be an inclusive method to enable people with knee OA to reap the benefits of high-impact exercise participation, without the risk of damaging cartilage or worsening pain.

## Introduction

Osteoarthritis (OA) is a degenerative joint disease which leads to pain, stiffness, and loss of mobility. Clinically, OA most commonly affects the knee ^1^. While all international guidelines recommend physical activity and exercise (henceforth termed ‘exercise’) as the first-line intervention for managing knee OA ^2^, a common theme across these guidelines is the emphasis on low-impact exercises. However, high-impact exercises like running, jumping, and hopping have many superior health benefits compared to low-impact exercises, such as providing a better mechanical stimulus to strengthen bones ^3, 4^. This would be beneficial for people with knee OA, as they are typically older and thus have an increased risk of osteoporosis.

Unfortunately, people with knee OA are more sedentary^5^ and physically more deconditioned ^6^ than age-matched healthy controls. They may also fear that exercise may worsen their knee pain ^7^. These factors are likely to deter people with knee OA from engaging in higher-impact exercises. Alternating between walking and running (herein termed ‘walk-run’) may be a more inclusive approach to encourage participation in high-impact exercises. Alternating between walking and running is routinely undertaken within a progressive training program (e.g. Couch to 5k) to help people transition from being sedentary to running 5 km. A previous study reported that one minute of high-impact daily exercise was positively associated with greater bone mineral density T-score in pre-menopausal women ^8^. This suggests that walk-run may provide similar benefits to continuous running for people with knee OA, with less risk of exacerbating pain and disease progression.

It is well-established that a single bout of exercise can have analgesic effects; this has been termed ‘exercise-induced hypoalgesia’ (EIH) ^9, 10^. EIH represents a temporary reduction in pain sensitivity immediately following exercise ^9^. For example, cycling between 4-10 min resulted in a 15% increase in pressure pain threshold (PPT) in healthy people ^11^. Additionally, greater exercise intensity evokes a stronger EIH response than a lower exercise intensity in healthy people ^10^. Higher intensity exercise could therefore be beneficial to those with knee OA. However, like those with other musculoskeletal pain conditions, people with knee OA have a more variable EIH response to exercise ^9, 11, 12^, which justifies further investigation.

Few studies have investigated the effects of high-impact exercises in knee OA. A prospective imaging study reported that running did not accelerate structural progression in people with knee OA ^13^. Another study reported a delayed increase in T2 relaxation times of the knee cartilage in people with knee OA compared to healthy controls after 30 minutes of running ^14^. A greater T2 relaxation time reflects a greater time needed for water to return to equilibrium in the knee cartilage, suggesting that people with knee OA may need more time to recover than healthy people after running ^14^. In contrast to imaging, blood-based biomarkers offer another method of understanding the effects of exercise on cartilage health. One common cartilage biomarker is cartilage oligomeric matrix protein (COMP) ^15^; a non-collagenous glycoprotein primarily found in cartilage, with a crucial role in cartilage structure and turnover. It is postulated that COMP is a mechanosensitive protein, fluctuates in response to mechanical load on the joint ^16^. For example, in healthy people, running for 30 minutes increased COMP immediately by 30% from baseline, whereas walking for the same duration led to only a 9.7% rise from baseline ^17, 18^.

Whether adding small volumes of high-impact running to a low-impact walking program will alter the pain and biological response differently to walking is unclear. Understanding this is essential to developing more inclusive exercise programs for people with knee OA that can mitigate age-related musculoskeletal deterioration, without exacerbating the disorder. This study aims to investigate the biological and pain responses to a walk-run program in people with self-reported knee OA. Given that COMP is mechanosensitive, we also aimed to measure local load environment by quantifying knee joint moments ^19, 20^, to better understand how the walk-run program alters cartilage metabolism. The present study has three hypotheses: first, COMP change from baseline would exceed 10%, which represents a level found in walking ^17, 18^; second, a walk-run program would induce an EIH response greater than an effect size (Hedge’s *g*) of 0.4 ^9^; lastly, COMP change from baseline would have a moderate positive correlation with cumulative knee joint moments.

## Methods

### Study Design

This was a cross-sectional study design involving a single-session laboratory testing. Participants aged 35–75 years with self-reported knee OA ^21^, pain intensity ≤ 3/10 on a verbal numerical rating scale, a body mass index of < 35 kg/m², the ability to ambulate independently, and good general health were eligible to participate. Participants with health conditions causing sensory deficits (e.g., diabetes), taking medication affecting sensation, currently pregnant, or with a history of chemotherapy or terminal illness were excluded. The study was approved by the University of Essex Ethics Sub-Committee 2 (ETH2324-1428).

### Self-reported outcome measures

Participants were asked to provide demographic information, including age, sex, dominant leg, the affected side of OA, and the duration of their condition. The Knee Injury and Osteoarthritis Outcome Score (KOOS) was administered to assess knee function and symptoms. The KOOS includes five subscales: Pain, Symptoms, Activities of Daily Living, Sports and Recreation Function (impact of knee issues on high-demand activities), and knee-related Quality of Life. Each subscale is scored on a scale from 0 to 100, where 100 indicates no symptoms and 0 represents extreme symptoms ^22^. Additionally, the UCLA Activity Score was used to evaluate participants’ physical activity levels, categorising them into low activity (1–4), moderate activity (5–7), and high activity (8–10) ^23^. The perceived knee pain of the affected limb was evaluated on an 11-point verbal numerical rating scale (0 no pain, 10 maximal tolerable pain) before and during the protocol at 8:30 minutes, 16:00 minutes, and 23:30 minutes.

### Walk-Run Protocol

Participants walked on a force-plate instrumented treadmill (Bertec, USA) at a self-selected comfortable speed, which was recorded, for five minutes as a warm-up, followed by a 20 minutes alternating bouts of jogging and walking: 60 seconds of jogging at their comfortable speed, which was recorded, interspersed with 90 seconds of walking at the recorded preferred speed (Figure 1). This walk-run protocol was used as it reflected the first session of the Couch to 5K program – a graded training program designed to prepare novice runners towards completing a 5km run. During this protocol, biomechanical data were collected at three time points (Figure 1).

**Figure 1.**
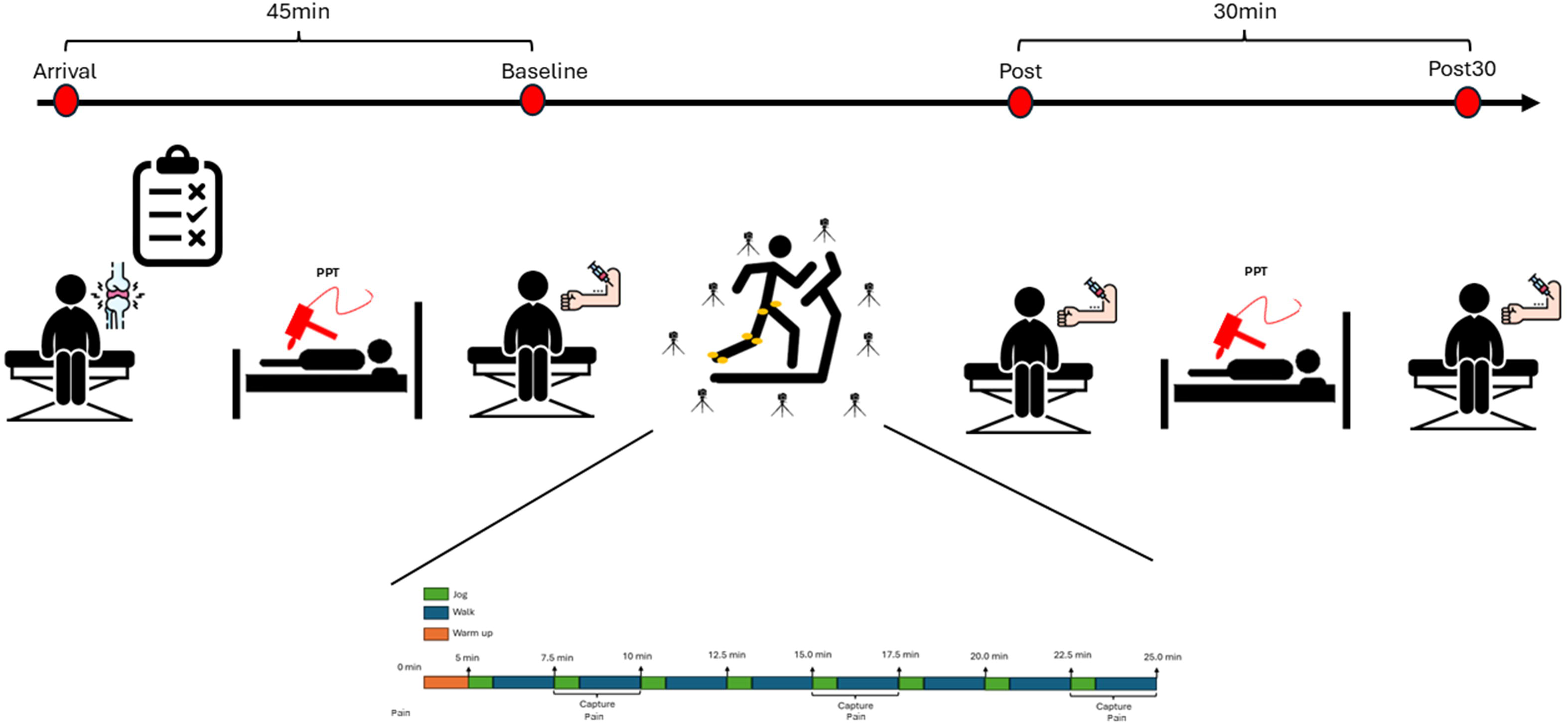
Schematic illustration of the data collection protocol.

### Biomechanics Modelling and Processing

Retroreflective markers (14 mm diameter) were placed on the pelvis (bilateral anterior and posterior superior iliac spines), bilateral medial and lateral femoral condyles and malleoli, the first and fifth metatarsals and mid-calcaneus of the affected side. In addition, rigid clusters of four markers were placed on the lateral thigh and shank of the affected side. Marker trajectories were captured at 200 Hz using a 9-camera motion analysis system (Vicon T-series, Oxford Metrics, UK), while GRF was recorded at 2000 Hz using a force plate treadmill. The position and orientation of the lower limb segments were calculated using an inverse kinematic (IK) lower limb model created in Visual 3D (HAS-motion, Germantown, MD). The hip joint centre was defined using a regression equation, whilst the knee and ankle joint centres were defined as the mid-point of the femoral epicondyles and malleoli, respectively. For the IK model, the hip, knee, and ankle joints were constrained to have three rotational degrees of freedom (DOF), whilst the pelvis segment had six DOF.

Raw marker trajectories and force data were filtered using a low-pass, zero-lag, 4th-order Butterworth filter at 18 Hz ^24^. Inverse dynamics analysis was then performed in Visual 3D to compute the three-dimensional (3D) internal knee joint moments, which were expressed in the proximal segment’s reference frame. Joint moments were normalized to body mass (Nm/kg). The knee resultant moment (scalar magnitude of three axes) of the target limb was calculated. Three biomechanical load indices were extracted for each participant. Peak knee resultant moment was quantified during the steady-state walking and running period, separately, and the average across all cycles was calculated for each. The cumulative knee resultant moment was calculated across the 25 minutes session. To do this, we first quantified the impulse of the absolute knee resultant moment across a complete stride of walking and running, separately. We then averaged the knee moment impulse values across all walks and all running cycles, separately. Next, we quantified the average steps per minute for both walking and running, separately. Since the participants undertook eight minutes of running and 17 minutes of walking, the total number of steps walked and run was calculated, multiplied by their respective per-stride impulse values, and summed to determine the cumulative impulse.

### Blood Sampling Protocol and Processing

Blood samples were obtained before commencement of the walk-run protocol (Baseline), then immediately (Post) and 30 min afterwards (Post30). Venous blood samples (6 ml) were obtained via venipuncture of the median cubital vein in the antecubital fossa at each time point. After clotting for 60 minutes at room temperature, samples were centrifuged for 15 minutes at 1000g at 6°C, before aliquoting into 1.5ml cryovials and storage at 70°C until further analysis. Human COMP concentration was determined using a commercially available enzyme-linked immunosorbent assay (Quantikine ELISA #DCMP0; R&D Systems, Minneapolis, MN). The assay has a sensitivity of 0.036 ng/mL and an assay detection range of 0.156 to 10 ng/mL, with inter- and intra-assay coefficients of variation < 5%. All the assays were run in duplicate. COMP (%) changes from post exercise (Post) and 30 minutes post exercise (Post30) relative to the baseline, were calculated using: 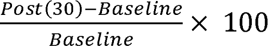.

### Pressure pain thresholds

PPTs were measured at the medial knee joint line of the affected knee and the head of the radius of the ipsilateral elbow, using a hand-held digital pressure algometer (Medoc Ltd, Israel) with a contact probe with an area of 1 cm^2^. The algometer probe was positioned perpendicular to the skin during testing. A loading rate of 30 kPa/s, based on real-time on-screen applied force feedback, was used ^25^. During PPT testing, participants remained in a supine position with legs extended. Participants were asked to press a button with their contralateral hand, indicated by an instructional cue: “Pressure will be applied at a gradual rate. On a scale of 0–10, where 0 is no pain and 10 is the pain as bad as it could be, allow the pressure to increase until it reaches a point where you begin to feel pain with an intensity of 2/10, and then press the button” ^25^. Two repetitions of PPT testing were completed at the knee and the elbow, and the average of the values at each site was calculated. If the participant failed to report pain at the level referred to within a given cue, the test would be stopped at an application of 1000 kPa pressure for safety purposes, with this value recorded as the PPT. A 1-min interval was observed between consecutive PPT assessments ^25^. PPTs were undertaken after 45 minutes of non-weightbearing rest, and approximately 15 minutes after the end of the walk-run program. A gap of 15 minutes enabled blood samples to be taken immediately after exercise. EIH was calculated using the formula:

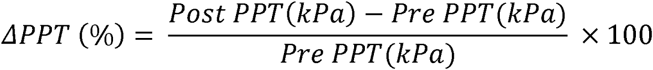

### Sample size

A previous study reported an effect size in COMP change from baseline of 0.9 (12.3%/13.2%) ^18^. Based on a one-sample t-test, with an alpha of 0.05 and a power of 0.8, a sample size of 12 was planned.

### Data Analysis

Statistical analysis was performed using R software (v4.4.2) software. Descriptive statistics, including mean ± standard deviation (SD), were reported for all numeric variables. For the primary outcome, COMP concentration changes from post-exercise to 30 minutes post-exercise were computed by 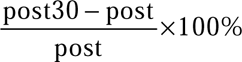. To explore the relationship between mechanical loading and COMP, Pearson correlation coefficients were computed between percentage changes in COMP and three biomechanical variables separately. Statistical significance was determined with an alpha of 0.05. Correlation coefficients were interpreted using commonly accepted thresholds in medical research, where *r* < 0.3 was considered a weak correlation, 0.3 ≤ *r* < 0.5 a moderate correlation, and *r* ≥ 0.5 a strong correlation ^26^.

## Result

Eight participants volunteered to participate within the window of data collection, and all took part in the study (Table 1). Participants walked at a mean speed of 0.96 (SD: 0.21) m/s and jogged at a mean speed of 1.64 (SD: 0.46) m/s during the walk-run task.

**Table 1:** Characteristics of the participants.

| Characteristics | Participant (*n* = 8) |
| --- | --- |
| Age, mean (SD), years | 52.0 (10.3) |
| Gender, *n* (%) |  |
| - Male | 6 (75%) |
| - Female | 2 (25 %) |
| Height, mean (SD), m | 1.77 (0.12) |
| Body mass, mean (SD), kg | 90.64 (17.3) |
| Leg of dominant, right leg, *n* (%) | 7 (87.5%) |
| Affected osteoarthritic side, *n* (%) |  |
| - Left | 2 (25%) |
| - Right | 4 (50%) |
| - Both legs | 2 (25%) |
| Duration of symptom, *n* (%) |  |
| - Less than 1 year | 1 (12.5%) |
| - 1-2 years | 2 (25%) |
| - 3-5 years | 2 (25%) |
| - More than 10 years | 3 (37.5%) |
| KOOS, mean (SD) |  |
| - Symptoms | 67.857 (13.0) |
| - Pain | 69.097 (11.1) |
| - ADL | 79.228 (9.0) |
| - Sports | 48.75 (17.5) |
| - QOL | 46.875 (15.6) |
| UCLA, *n* (%) |  |
| - Regularly participates in impact sports | 2 (25%) |
| - Regularly participates in moderate activities | 2 (25%) |
| - Sometimes participates in moderate activities | 2 (25%) |
| - Sometimes participates in impact sports | 2 (25%) |

Figure 2 reports the absolute and percentage change of COMP at baseline, immediately after exercise, and 30 minutes after exercise. The percentage change of COMP significantly increased from baseline immediately after exercise (*t* = 2.29, P = 0.028, Hedges’ *g* = 1.43 [95%CI 0.46, 2.37]), then returned to baseline concentration at Post30 with no significant difference between the two time-points (t = 0.21, P = 0.419, Hedges’ *g* = 0.07 [95%CI −0.55, 0.68]) (Figure 2).

**Figure 2.**
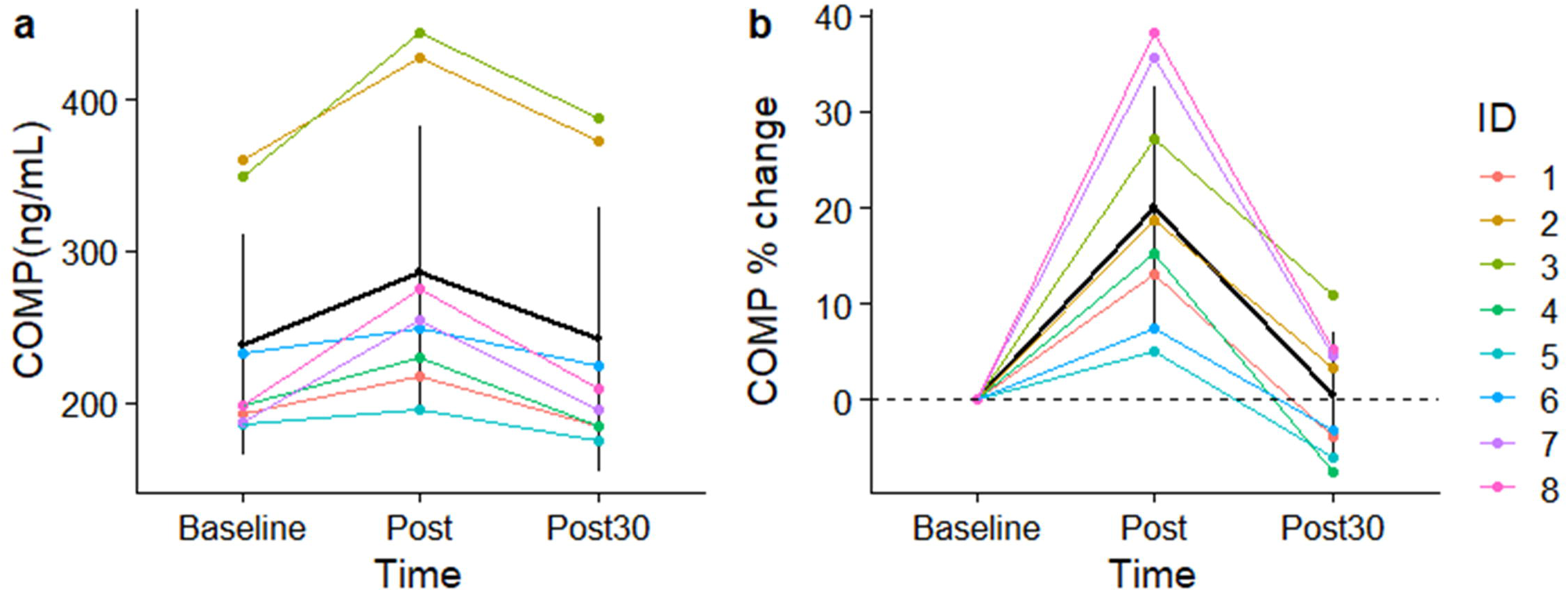
(a) Absolute levels and (b) relative increases in Cartilage Oligomeric Matrix Protein (COMP) levels at each of the three time points. Black points indicate the mean with error bars representing one standard deviation.

PPT did not significantly change from baseline at the knee (LJPPT = 7.28% [95%CI −35.63, 50.19%], *t* = 0.40, *P* = 0.700) and at the elbow (LJPPT = −9.15% [95%CI −39.28, 20.97%], *t* = −0.72, *P* = 0.496) (Figure 3). This corresponded to a Hedges’ *g* = 0.13 (95%CI −0.50, 0.74) and *g* = −0.23 (95%CI −0.84,0.41) for the knee and elbow, respectively. Perceived knee pain intensity was 0.4(0.7), 1.0 (0.9), 1.4 (1.3), and 1.3 (1.4), before the protocol, and at 8:30 minutes, 16:00 minutes, and 23:30 minutes, respectively (Figure 3c).

**Figure 3.**
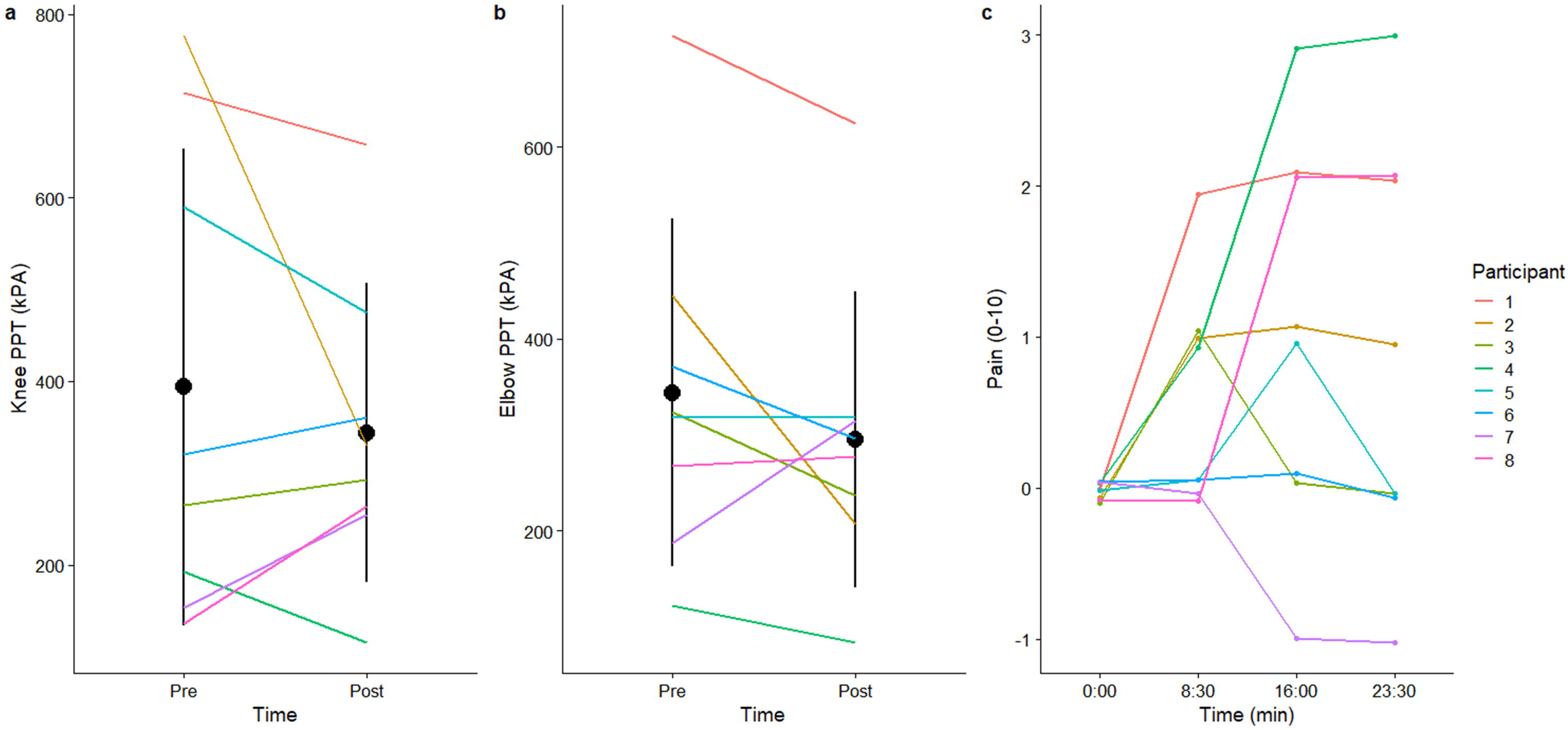
Absolute levels of pressure pain threshold (PPT) values of the (a) knee, and (b) elbow; and (c) self-reported pain. Black points indicate the mean with error bars representing one standard deviation.

Figure 4 illustrates the time-series of the knee resultant moment of a representative subject and the segmented knee moments across a single walking and running stride. The participants walked and jogged with an average peak knee resultant moment of 0.76 (0.13) Nm/kg and 1.87 (0.53) Nm/kg, respectively. Across the exercise, participants accumulated a knee resultant moment impulse of 484.66 (52.74) Nm.s/kg. No statistically significant associations existed between the knee load indices and COMP alterations (Table 2). However, four out of six comparisons achieved a moderate effect size but were not statistically significant (Table 2). Figure 5b also reports the results of two simple simulations – walking for 25 minutes and jogging for 25 minutes. The cumulative knee impulse is 387.85 (52.38) Nm.s/kg and 690.37 (124.73) Nm.s/kg for 25 minutes walking, and 25 minutes running, respectively. This means that the cumulative knee impulse was 20% lower for pure walking, and 42% greater for pure running, than walk-run.

**Figure 4.**
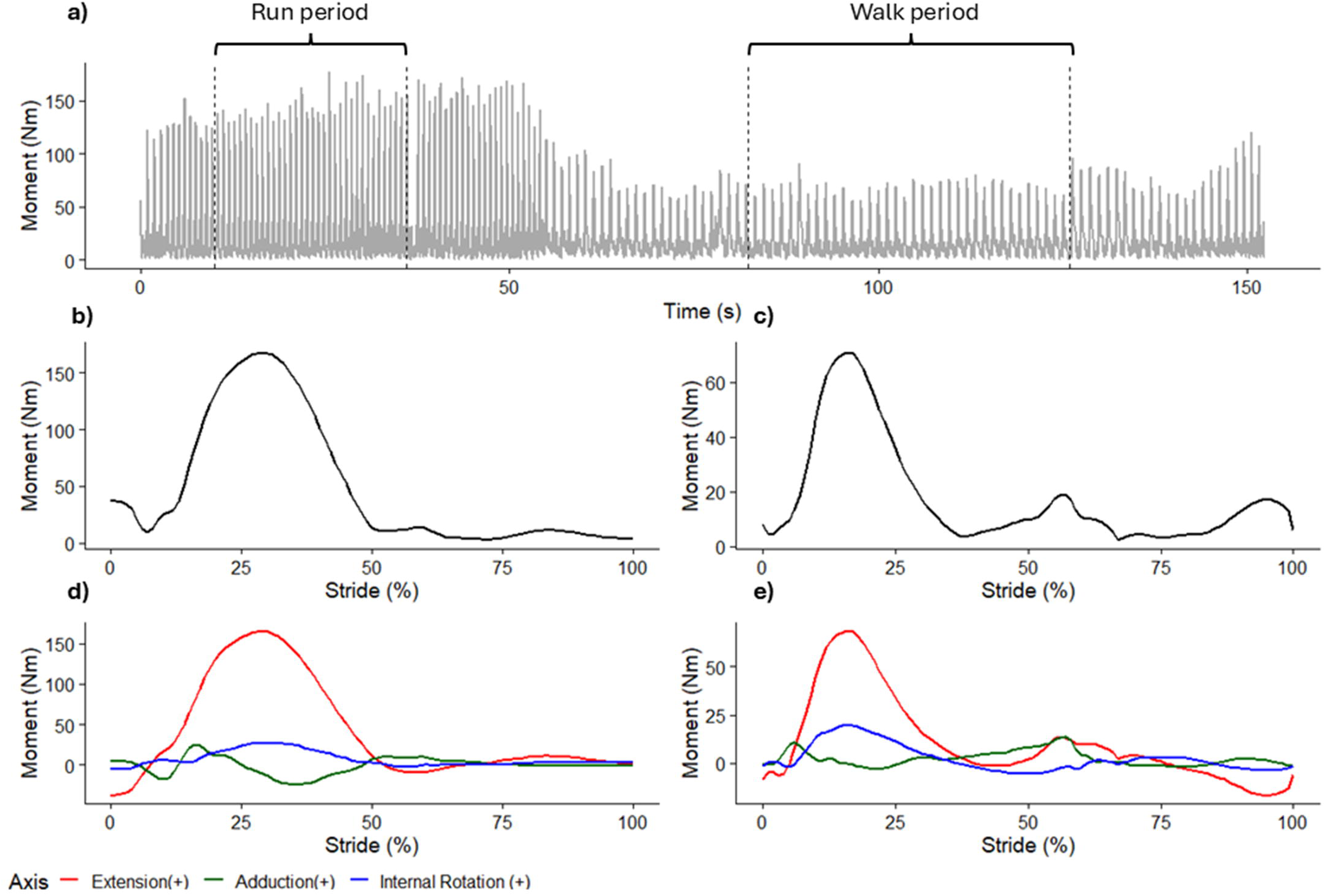
(a) An example of a continuous time-series of one subject (Subject 7)’s knee resultant joint moment. An example of a time-normalised waveform of the same subject’s knee resultant moment during running (b), and walking (c). An example of a time-normalised waveform of the same subject’s three-dimensional knee moments during running (e), and walking (f).

**Table 2.** Association between different load indices and COMP alterations.

| **COMP** | **Load** | **r** | **LB** | **UB** | **t** | **P value** |
| --- | --- | --- | --- | --- | --- | --- |
| **Post** | Cumulative | -0.22 | -0.80 | 0.58 | -0.546 | 0.605 |
| **Post** | Peak walk | 0.38 | -0.44 | 0.86 | 1.002 | 0.355 |
| **Post** | Peak run | -0.45 | -0.88 | 0.37 | -1.248 | 0.259 |
| **Post30** | Cumulative | -0.40 | -0.86 | 0.42 | -1.078 | 0.322 |
| **Post30** | Peak walk | -0.19 | -0.79 | 0.60 | -0.469 | 0.656 |
| **Post30** | Peak run | -0.33 | -0.84 | 0.49 | -0.844 | 0.431 |

**Abbreviations:** COMP - Cartilage Oligomeric Matrix Protein; r – Pearson correlation magnitude; LB – lower bound of 95% confidence interval, UB – upper bound of 95% confidence interval; t – t value

## Discussion

This represents the first study to simultaneously investigate the biological, pain, and biomechanical effects of a more inclusive method of introducing small volumes of high-impact exercise in people with knee OA. We found that walk-run resulted in an immediate increase in COMP concentration after exercise, exceeding changes typically associated with a walking-specific program. However, a rapid return of COMP to baseline suggests that the mechanobiological effects of walk-run did not reach the level of pure running. Contrary to our hypotheses, our walk-run program did not alter pain sensitivity.

The lack of EIH response was not surprising given that reported magnitudes and directions of alterations in pain sensitivity in musculoskeletal pain conditions are known to be highly variable ^9^. A recent study reported that those with knee OA who experienced an increase in perceived pain during exercise were more likely to experience EIH after exercise ^27^. Specifically, a 1-point (out of 10) increase in pain during exercise increased the odds of experiencing EIH by 43% ^27^. However, our results do not support this observation. For example, Participant 4 experienced an increase in perceived pain by 2/10 but did not exhibit an increase in PPT values after exercise (Figure 3). Also, Participant 7 experienced less pain during exercise, but exhibited an increase in PPT values after exercise (Figure 3). Another study reported that subgroups of knee OA patients with abnormal conditioned pain modulation (CPM; i.e. no pain inhibition mechanism after a second, noxious stimulus), are less likely to experience EIH, compared to those with normal CPM response ^28^. Future studies to investigate if there are subgroups of people with knee OA who would experience greater EIH from engaging in higher-impact exercises are warranted.

Despite the absence of a significant EIH response, walk-run did not increase pain sensitivity either. An increase in pain sensitivity with exercise may interfere with long-term exercise engagement and adherence. By its definition, high-impact exercises create more knee tissue load than low-impact exercises ^29^, and can activate more nociceptors, thereby triggering more pain. The present finding is supported by another study on people with back pain, which found that high-intensity aerobic exercises did not increase pain sensitivity, compared to moderate-intensity exercises ^30^.

Changes in COMP concentration provide evidence that the walk-run induced a greater biological cartilage response than a pure walking program, but not to the level of a pure running program. Post COMP increased by 20% in the present study, whereas it has been shown to increase by <10% in walking ^17, 31, 32^ By comparison, Post COMP has been shown to increase by 29 to 39% immediately following running ^33–35^. COMP has been shown to return to baseline within 30 minutes after a 30-minute walk ^17^, compared to 60 minutes after a 30-minute run ^36^. Yet, we found that Post30 COMP returned to baseline after the walk-run. These differences are likely because the cumulative knee joint load of walk-run was only 20% greater in a similar duration walking program, but 42% lower in a similar duration running program.

Surprisingly, COMP associations with all knee joint moment indices did not reach statistical significance, even though some associations exceeded the correlation threshold to be considered moderate in magnitude. An immediate observation could be that a lack of significance could be attributed to a small study sample size. However, previous studies did not report a significant association between COMP changes immediately after exercise with knee joint moments ^20, 37^. Unlike prior studies focusing only on per-step moment indices ^20, 37^, the present study quantified the cumulative moment impulse, which we anticipated to provide a more accurate knee load profile. We calculated joint moments using inverse dynamics, which does not consider differences in muscle activation patterns. Kersting et al. (2005) reported that knee muscular co-activation was the main mechanical parameter related to cartilage volume changes after running and not knee joint moments. Given that muscle forces are the greatest contributor to joint loads, future research should investigate the effects of neuromuscular indices during motor activities and their association with cartilage biomarkers.

This study is not without limitations. First, we did not achieve our target sample size of 12. However, prior research has also employed small cohorts of 5 to 10 participants when studying blood biomarkers in people with OA ^17, 35^. Second, we allocated all participants to a single group, which precluded comparing our results with another exercise condition. However, the plethora of studies which have investigated COMP and EIH responses in simpler walking or running conditions enables us to make comparative inferences in our statistical analysis. Third, we included people with self-reported knee OA, using a validated questionnaire, rather than people with an objective medical diagnosis coupled with imaging. Fourth, unlike previous studies that used continuous running ^36^, our protocol introduced frequent transitions between acceleration and deceleration phases. Positive and negative knee joint work is 33% and 41% lower in acceleration running than steady-state running, respectively ^38^. We did not use a continuous capture of the biomechanics data over 25 minutes because of the intensive computational processing power and memory required. With rapid advancement in wearable sensors and machine learning, it may soon be possible for continuous, prolonged analysis of knee joint loads ^39^. Lastly, we did not quantify the potential for a delayed increase in COMP up to 5 hours post-exercise ^17^. Previous studies have reported that OA patients with higher COMP levels 3.5 and 5.5 h after a 30_min walking exercise, relative to baseline resting values, demonstrated greater cartilage thinning over 5 years ^40^.

## Conclusion

Including short periods of running within a primarily walking-based session does not lead to prolonged elevations in cartilage stress markers and altered pain sensitivity. Taken together, these findings provide preliminary evidence that this interval approach to exercise may be a viable, low-barrier option for people with knee OA who are hesitant to engage in higher-impact exercises due to fear of pain and accelerating structural progression of the disorder.

## Data Availability

All data produced in the present work are contained in the manuscript

## Author contributions

Conceptualization (BL, CM, NU, SW, DE), Methodology (BL, CM, NU, SW, DE), Data curation (TC, BL, NU, CM, DE), Supervision (BL, DE, SW, CM), Data Processing and Analysis (TC, NU, CM, QM, BL), Validation (All authors), Writing-Original draft preparation (All authors), Writing – Reviewing and Editing (All authors).

## Acknowledgements

We would like to thank Jasmine Carvajal Cortes and Daniel Sporle for helping with data collection.

## Sources of funding

Dr Liew is supported by the Medical Research Council (MR/Y013557/1) and Innovate UK (Ref: 10093679). The funder had no role in the design, data collection, data analysis, and reporting of this study.

## Conflict of Interest

The authors have no conflicts of interest to declare.

## Notes

### Competing Interest Statement

The authors have declared no competing interest.

### Author Declarations

The study was approved by the University of Essex Ethics Sub-Committee 2 (ETH2324-1428).

